# Safety and immunogenicity of heterologous and homologous inactivated and adenoviral-vectored COVID-19 vaccines in healthy adults

**DOI:** 10.1101/2021.11.04.21265908

**Authors:** Nasamon Wanlapakorn, Nungruthai Suntronwong, Harit Phowatthanasathian, Ritthideach Yorsaeng, Preeyaporn Vichaiwattana, Thanunrat Thongmee, Chompoonut Auphimai, Donchida Srimuan, Thaksaporn Thatsanatorn, Suvichada Assawakosri, Sitthichai Kanokudom, Natthinee Sudhinaraset, Yong Poovorawan

**Affiliations:** Center of Excellence in Clinical Virology, Faculty of Medicine, Chulalongkorn University, Bangkok, 10330, Thailand; FRS(T), the Royal Society of Thailand, Sanam Sueapa, Dusit, Bangkok 10330, Thailand

**Keywords:** heterologous, homologous, inactivated, COVID-19, vaccine, viral vectored, adults

## Abstract

In light of intermittent supply shortages of individual vaccines and evidence of rare but serious adverse events after vaccination, heterologous regimens for COVID-19 vaccines have gained significant interest. This study aims to assess the reactogenicity and immunogenicity of the heterologous adenoviral vector vaccine regimen (ChAdOx1-S, AstraZeneca; hereafter referred to as AZ) and the inactivated vaccine regimen (CoronaVac; hereafter referred to as CV) regimen in healthy Thai adults immunized between June and September 2021. Our study showed that adverse events following homologous CV-CV and AZ-AZ, and heterologous CV-AZ and AZ-CV combinations, were mild and well tolerated overall. Receptor-binding domain (RBD)-specific antibody responses and neutralizing activities against wild-type and variants of concern after two-dose vaccination were higher in the heterologous CV-AZ and homologous AZ-AZ groups compared to the CV-CV and AZ-CV groups. Conversely, the spike-specific IgA response was detected only in the CV-AZ group after two doses of vaccination. The total interferon gamma response was detected in both the CV-AZ and AZ-CV groups after the two-dose vaccination. Given the shorter completion time of two doses, heterologous CoronaVac followed by ChAdOx1-S can be considered as an alternative regimen to homologous efficacy-proven ChAdOx1-S in countries with circulating variants. Additional studies on the efficacy and durability of immune responses induced by heterologous vaccine regimens are warranted.

## Introduction

In light of intermittent supply shortages of individual vaccines and evidence of rare, but severe, adverse events following vaccination, heterologous regimens for COVID-19 vaccines have gained significant interest. Several European countries have recommended heterologous messenger RNA (mRNA) boost strategies for people aged 55 to 65 years who have received one dose of adenoviral vectored vaccine (ChAdOx1-S, AstraZeneca, Oxford, UK) (AZ) due to concern about thrombotic events after a second ChAdOx1-S vaccination.^1^ Randomized and observational studies have demonstrated a potent induction of severe acute respiratory syndrome coronavirus 2 (SARS-CoV-2) S-specific antibodies and T-cell responses after a heterologous boost with the mRNA vaccine in individuals primed with the ChAdOx1-S vaccine.^2-5^ Furthermore, in ChAdOx1-S-primed individuals, a ChAdOx1-S boost did not induce potent neutralizing antibodies against the delta variant compared to a heterologous boost with an mRNA vaccine.^6^

Other mix and match vaccine strategies have been reported in addition to adenoviral vectored/mRNA vaccines. The heterologous prime boost trial of AstraZeneca ChAdOx1-S and inactivated whole virion BBV152 (Covaxin) in India found that immunization with adenoviral vector vaccine followed by inactivated whole virus vaccine was safe and elicited better immunogenicity than the homologous whole virus inactivated vaccine.^7^ Another study in China showed that the administration of the COVID-19 recombinant adenovirus type-5 vectored vaccine (Convidecia) as a second dose in individuals primed with inactivated CoronaVac induced better antibody responses than CoronaVac two-dose, with an acceptable reactogenicity profile.^8^

Between March and July 2021, Thailand had only two COVID-19 vaccines available: the inactivated and ChAdOx1-S from AstraZeneca. The CoronaVac vaccine was associated with ‘immunization stress-related responses (ISRR)’ characterized by numbness, or sometimes weakness, in the limbs among healthcare workers prioritized for CoronaVac.^9-10^ Although this self-limited adverse event is rare, people experiencing this side effect sought the ChAdOx1-S regimen for their second shot. Based on preliminary immunogenicity data for CoronaVac followed by the ChAdOx1-S vaccine from individuals seeking blood tests^11^ and the inadequate supply of the second dose of CoronaVac, the Ministry of Public Health of Thailand announced on 12 July 2021 that CoronaVac followed by ChAdOx1-S 3 to 4 weeks apart as an alternative regimen for healthy Thai people and those experiencing adverse effects after CoronaVac vaccination.^12^ This study was designed and conducted before the official announcement of the CoronaVac/ChadOx1-S regimen. Our aim was to evaluate the safety and immunogenicity of heterologous CoronaVac immunizations followed by the ChAdOx1-S and ChAdOx1-S followed by CoronaVac in a prospective cohort of healthy Thai adults. The results of this study will help guide the physician’s decision on a mix-and-match vaccine strategy in certain circumstances, such as vaccine shortages or adverse events after vaccination.

## Patients and Methods

### Study cohort

This was a prospective cohort study that enrolled healthy Thai adults aged ≥18 years. A total of 180 immunocompetent individuals immunized with the homologous inactivated COVID-19 vaccine (CoronaVac; hereafter referred to as CV) (n=90) and the adenoviral vector vaccine (ChAdOx1-S, AstraZeneca; hereafter referred to as AZ) (n=90) were previously enrolled between March and May 2021 (Thai Clinical Trials Registry; TCTR20210319003). A total of 46 and 48 immunocompetent individuals were prospectively enrolled to receive heterologous CV followed by the AZ (CV-AZ group) and the AZ followed by the CV vaccines (AZ-CV group) group, respectively (TCTR20210628005) between June and September 2021. The inclusion criteria were immunocompetent individuals older than 18 years of age with no or well-controlled comorbidities and no previous SARS-CoV-2 infection from the medical history. The study flow showing the enrollment of participants and the sample size in this study is demonstrated in Figure S1. The study protocols were approved by the Institutional Review Board (IRB) of the Chulalongkorn University Faculty of Medicine (IRB numbers 192/64 and 491/64). Written informed consent was obtained from all participants prior to collecting clinical data and samples. This study is conducted in accordance with the Helsinki Declaration of 1975.

### Vaccination and blood collection

CoronaVac (Sinovac Life Sciences, Beijing, China) is an inactivated virus vaccine created from African green monkey kidney cells (Vero cells) that have been inoculated with SARS-CoV-2 (CZ02 strain). At the end of the incubation period, the virus was harvested, inactivated with β-propiolactone and formaldehyde, concentrated, purified, and finally absorbed into aluminum hydroxide. Each vial contains 0.5 mL with 600 Spike Units (equal to 3 microgram) of inactivated SARS-CoV-2 whole virus as antigen.^13^ CoronaVac was approved for Thai adults ages 18-59 and was administered 21-28 days apart.

Chimpanzee adenovirus Oxford 1 (ChAdOx1)-vectored vaccine (AZ) from Oxford/AstraZeneca is a non-replicating viral vector vaccine that stimulates an immune response against the SARS-CoV-2 spike protein. One dose (0.5 mL) contains no less than 2.5 × 10^8^ infectious units of chimpanzee adenovirus encoding the SARS-CoV-2 spike glycoprotein.^14^ AZ vaccination was administered at 10-week intervals. The first heterologous cohort received CV followed by AZ at an interval of 4 weeks. The second heterologous cohort received AZ followed by CV at a 10 week interval. Blood samples were collected before the first vaccination (pre-prime), before the second vaccination (pre-boost), and 4 weeks after the second vaccination (post-boost).

### Safety assessment

Participants recorded both local and systemic adverse events (AE) after immunization within 7 days as described previously^15^ using self-administered online and paper questionnaires. Explanation about data collection was given to participants by trained investigators during the vaccination visit.

### Antibody assays

Serum samples were evaluated for total immunoglobulins (Ig) specific to the receptor-binding domain (RBD) of the SARS-CoV-2 spike (S) protein using Elecsys SARS-CoV-2 S according to the manufacturer’s instruction (Roche Diagnostics, Basel, Switzerland). Anti-RBD IgG were tested using the SARS-CoV-2 IgG II Quant assay (Abbott Diagnostics, Abbott Park, IL) according to the manufacturer’s instructions. Multiplying the numerical AU/mL by 0.142 converts the concentration to binding antibody units per milliliter (BAU/mL).

SARS-CoV-2 antinucleocapsid (anti-N) IgG was tested using the commercially available automated ARCHITECT system (Abbott Diagnostics, Abbott Park, IL) by chemiluminescent microparticle immunoassay (CMIA). The results are reported by dividing the sample result by the stored calibrator result. The default unit for the SARS-CoV-2 anti-N IgG assay is a sample/cut-off index (S/C). For interpretation, S/C ≥1.4 was defined as positive and S/C <1.4 as negative.

Anti-spike protein 1 (S1) IgA was tested using an enzyme-linked immunosorbent assay (ELISA) (Euroimmun, Lübeck, Germany). The results were derived from the ratio of optical density (OD) obtained from samples and the calibrator (CO). The maximum cutoff ratio (OD/CO) was 9; results >9 were recorded as 9.

The neutralization activity of the samples against wild-type SARS-CoV-2 and variants of concern B.1.1.7 (alpha), B.1.351 (beta), and B.1.617.2 (delta) were also evaluated using an ELISA-based surrogate virus neutralization test (sVNT). NeutraLISA (Euroimmun, Lübeck, Germany) was used to evaluate the wild-type strain only. The cPass SARS-CoV-2 neutralizing antibody detection kit (GenScript, Piscataway, NJ) was used for all strains. The recombinant RBD from B.1.1.7 (containing N501Y), B.1.351 (containing N501Y, E484K, and K417N), and B.1.617.2 (containing L452R and T478K) were used with this kit. Briefly, serum samples were diluted 1:10 with buffer and incubated with horseradish peroxidase conjugated RBD for 30 min at 37°C. Next, 100 µL of the sample mixture was added to a capture plate pre-coated with human angiotensin-converting enzyme 2 (ACE2) and incubated for 15 min at 37°C. After washing, 100 µL of TMB chromogen solution was added and the plate was incubated in the dark for 15 min at room temperature. After the addition of a 50 µL stop solution, the samples were read at 450 nm. The ability of a serum to inhibit binding between RBD and ACE2 was calculated as percentage as follows: 1 – (average OD of sample/average OD of negative control), multiplied by 100.

### Interferon gamma release assay

SARS-CoV-2-specific T cell responses were evaluated using a whole blood interferon-gamma release assay (IGRA); QuantiFERON (QFN) SARS-SoV-2 assay (Qiagen, Hilden, Germany). This assay uses two sets of SARS-CoV-2 spike (S) protein (Ag1 and Ag2). The SARS-CoV-2 Ag1 tube contains CD4+ epitopes derived from the S1 subunit of the spike protein. The Ag2 tube contains CD4+ and CD8+ epitopes from the S1 and S2 subunits of the spike protein. Briefly, heparinized blood samples were placed in Quantiferon® tubes containing spike peptides, as well as positive and negative controls. Whole blood was incubated at 37°C for 24 hours and centrifuged to separate plasma. interferon (IFN)-γ (IU/mL) was measured in these plasma samples using ELISA (QuantiFERON Human IFN-γ SARS-CoV-2, Qiagen) tests. IFN-γ values were subtracted from the unstimulated control (Nil) to mitigate against the background IFN-γ in the sample that was not the result of SARS-CoV-2 specific T cell stimulation. The detection limit of the test was 0.065 IU/mL. Typically, this assay used an 8-point standard curve, therefore, concentrations ≥ 10 IU/mL was defined as IU/mL.

### Statistical analysis

Baseline characteristics were reported as mean and range. RBD-specific total Ig and anti-RBD IgG were presented as geometric mean titers (GMT) with a 95% confidence interval (CI). Other parameters were presented in median with interquartile range. Differences in antibody titers, S/C, OD/CO, and percentage inhibition and IU/mL minus nil between groups were calculated using the Kruskal–Wallis test or the Wilcoxon signed rank test (nonparametric) with multiple comparison adjustments. The graphical presentations were prepared using GraphPad Prism version 9.0 software (Graph-Pad Software, Inc., San Diego, CA). The statistical analysis was performed using IBM SPSS Statistics for Windows, version 21 (IBM Corp., Armonk, NY). A p value <0.05 was considered statistically significant.

## Results

### Demographic data

The baseline demographic characteristics of the four groups of participants who received heterologous and homologous CV and AZ vaccines were similar (Table 1). The mean age of the participants in the CV-AZ, AZ-CV, CV-CV, and CV-AZ groups was 41.4, 43.1, 42.6, and 47.6 years, respectively. The median durations between the completion of the two-dose vaccination and blood sampling were between 30 and 32 days among all groups. In the CV-AZ group, there were 2 participants who were lost to follow-up; one was infected with SARS-CoV-2 before the booster dose was given and the other was unwilling to participate. In the AZ-CV group, there were 2 participants who were unwilling to receive the second dose of CoronaVac.

**Table 1.** Demographics and characteristics of the vaccinated cohorts.

|  | **CV-AZ** | **AZ-CV** | **CV-CV** | **AZ-AZ** |
| --- | --- | --- | --- | --- |
| n | 46 | 48 | 90 | 90 |
| Mean (range)<br>age, years | 41.1<br>(18.0-64.0) | 43.1<br>(23.0-59.0) | 42.6<br>(24.0-59.0) | 47.6<br>(19.0-85.0) |
| Sex |  |  |  |  |
| Male (%) | 26 (56.5) | 21 (43.8) | 49 (54.4) | 40 (44.4) |
| Female (%) | 20 (43.5) | 27 (56.2) | 41 (45.6) | 50 (55.6) |
| Time interval between first and second doses |  |  |  |  |
| Median<br>(range), days | 27.0<br>(27.0 -28.0) | 70.0<br>(70.0 -71.0) | 26.0<br>(21.0 -28.0) | 70.0<br>(70.0 -73.0) |
| Time interval between the second dose and blood sampling |  |  |  |  |
| Median<br>(range), days | 31.0<br>(30.0-35.0) | 32.0<br>(30.0-35.0) | 31.0<br>(28.0-49.0) | 30.0<br>(22.0-41.0) |

### Reactogenicity data of participants receiving the homologous and heterologous vaccine regimen

The most common solicited local adverse reaction (AE) after the first and second dose was pain at the injection site: CV-AZ group (first dose 42%; second dose 91%), AZ-CV group (first dose 54%; second dose 77%). The most common systemic AE was fatigue. The reported fatigue frequency was 25% and 82% in the CV-AZ group after the first and second dose, respectively, and 38% and 52% in the AZ-CV group after the first and second dose, respectively. Comparisons of AEs between first and second dose vaccination showed that local and systemic AEs after the first dose were higher than those reported after the second dose in the AZ-AZ group (Figure S2). On the contrary, the local and systemic AEs after the first and second doses of CoronaVac in the CV-CV group were not different. In addition, the heterologous group had a higher percentage of adverse events compared to the homologous group after the booster vaccination. (Figure 1). Most of the solicited local and systemic AEs requested were mild (grade 1) or moderate (grade 2) and resolved within a few days post-vaccination (Figure S2). Frequencies of grade 3 local or systemic AEs after the booster doses ranged from 2% to 5%. No serious AEs were reported.

**Figure 1.**
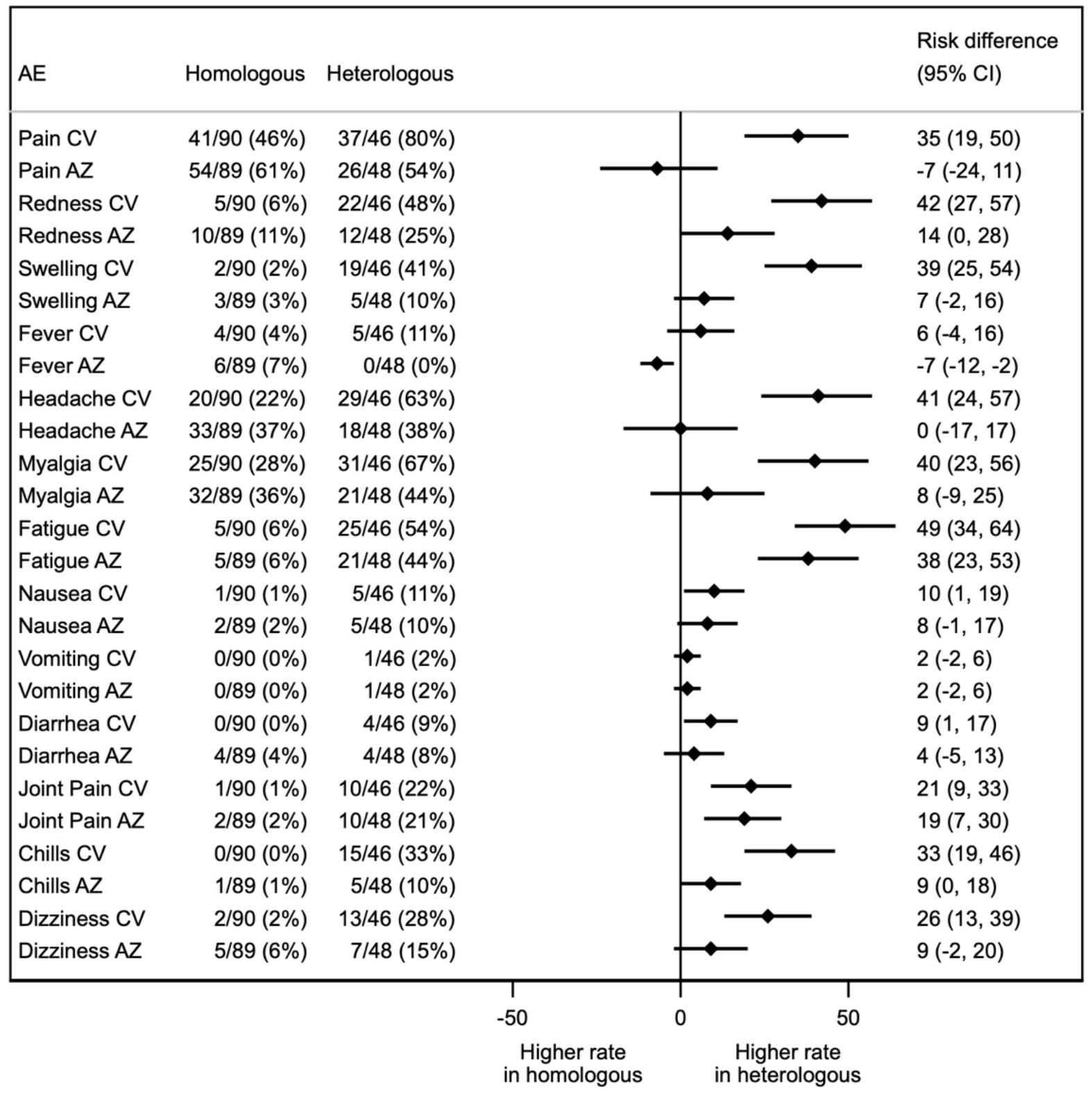
Forest plot showing the percentages of solicited local and systemic adverse events (AEs) and the absolute differences in the proportion of participants with any grade solicited AEs across 7 days post-boost vaccination with 95% confidence intervals. CV and AZ in the first column refer to the prime vaccination. AE denotes an adverse event.

**Figure 2.**
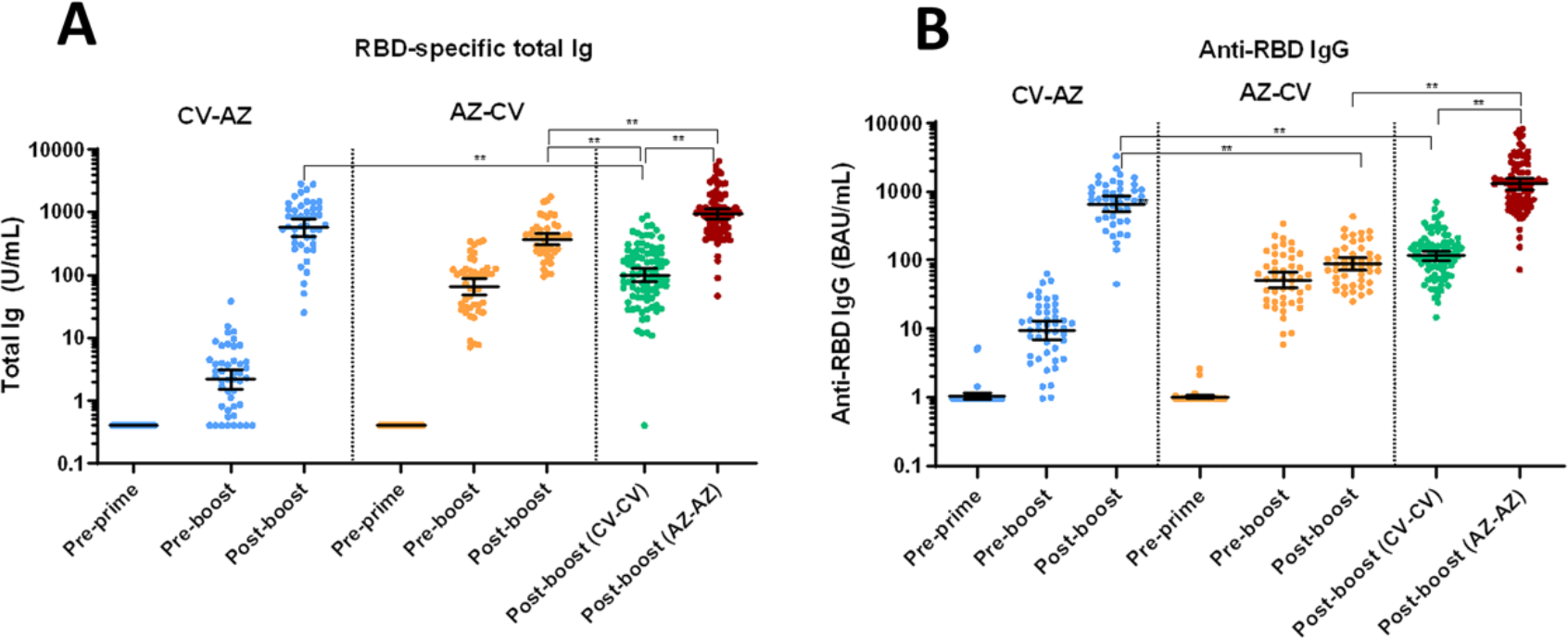
Binding antibody specific for SARS-CoV-2. A) Total immunoglobulin specific to the receptor-binding domain (RBD) (Ig) and B) Anti-RBD IgG in heterologous prime boost in CV-AZ and AZ-CV groups on the day of the first dose (pre-prime), 4 and 10 weeks later for CV-AZ and AZ-CV groups, respectively (pre-boost), and 4 weeks after two dose completion (post-boost). Data points are the reciprocals of the individual. Lines indicate geometric means and bars indicate 95% confidence intervals. As a reference, RBD-specific total Ig and anti-RBD IgG were compared 4 weeks after two-dose completion (post-boost) among homologous CV and AZ vaccinees. ** indicates *p* < 0.001.

### RBD-specific binding antibody after homologous and heterologous prime-boost *regimen*

The geometric mean titer (GMT) of total RBD immunoglobulin (Ig), comprised mainly IgG, IgM and IgA, and anti-RBD IgG were compared among all groups using the Kruskal–Wallis test with multiple comparison adjustment (Figure 1). The GMTs of the total RBD-Ig and anti-RBD IgG achieved after two doses of vaccination were similar between the CV-AZ group and the AZ-AZ group. The anti-RBD IgG levels achieved by the CV-AZ and AZ-AZ groups were significantly higher than those achieved by the CV-CV and AZ-CV groups (p <0.001). Interestingly, recipients of AZ following first dose CV vaccination had their anti-RBD IgG levels rise 70-fold in at one month after vaccination, compared with only a 13-fold increase in recipients of two-dose CV (Table S1).

AZ-primed individuals possessed higher total RBD Ig and anti-RBD IgG than CV-primed individuals in the pre-boost period. However, after CV enhancement, AZ-primed individuals had their anti-RBD IgG levels and total RBD Ig increased only 2-fold and 6-fold respectively, one month after vaccination, compared to a 27-fold and 14-fold increase in recipients of the two-dose AZ.

### Anti-N IgG and Anti-S1 IgA

The seropositivity rate of anti-N IgG was shown in Figure 3A. At baseline, none of the participants was positive for anti-N IgG. After two-dose vaccination, the anti-N IgG seropositivity rate was highest among the two-dose CV recipients. This can be explained by the fact that only the CV vaccine had the SARS-CoV-2 nucleocapsid protein. Our findings also demonstrated a significant increase in anti-N IgG only after a two-dose CoronaVac.

**Figure 3.**
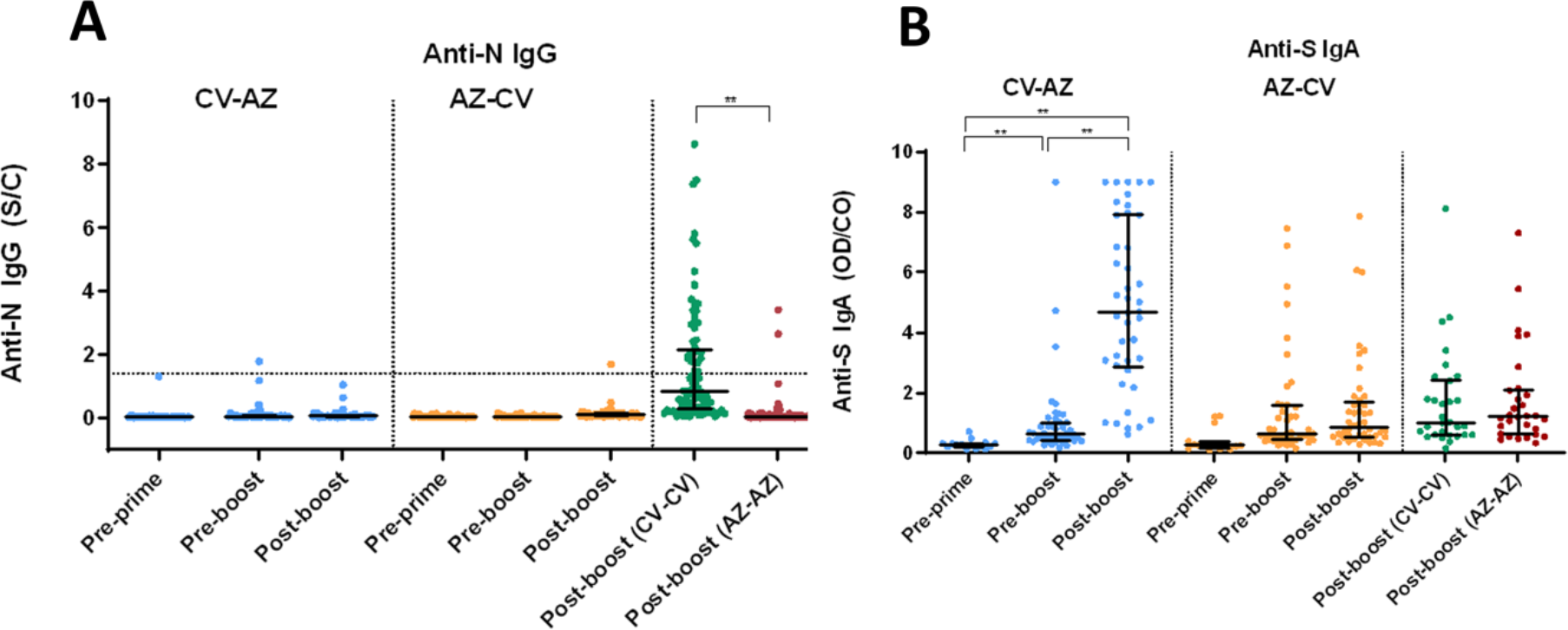
Anti-N IgG and Anti-S1 IgA. A) SARS-CoV2-specific nucleocapsid (N) IgG (Anti-N IgG) and B) Spike protein 1-specific IgA (anti-S1 IgA) in heterologous prime-boost CV-AZ and AZ-CV groups on the day of the first dose (pre-prime), 4 and 10 weeks later for CV-AZ and AZ-CV groups, respectively (pre-boost), and 4 weeks after two-dose completion (post-boost). The data points are the reciprocals of the individuals. Lines indicate median and bars indicate interquartile ranges. As a reference, anti-N IgG and anti-S1 IgA were compared 4 weeks after completion of the two doses (post-boost) among homologous CV and AZ vaccinees. ** indicates *p* < 0.001.

None of the participants was positive for serum anti-S1 IgA at baseline (Figure 3B). After the first dose of CV vaccination in the CV-AZ group, there was an increase in the OD/CO of anti-S1 IgA before the boost. Anti-S1 IgA OD/CO increased significantly only after two doses of CV followed by AZ.

### Neutralizing activities specific to SARS-CoV-2 wild-type and variant of concerns

NeutraLISA-specific neutralizing activities for wild-type SARS-CoV-2 after the first dose vaccination were higher in AZ-primed individuals than in CV-primed individuals (Figure 4). This finding agrees with the total RBD Ig and the anti-RBD IgG. However, following the second dose vaccination, the heterologous CV-AZ group possessed the highest neutralizing activities of all the groups (p < 0.001). There were also no significant differences in neutralizing activities after the second vaccination between the AZ-CV and CV-CV groups.

**Figure 4.**
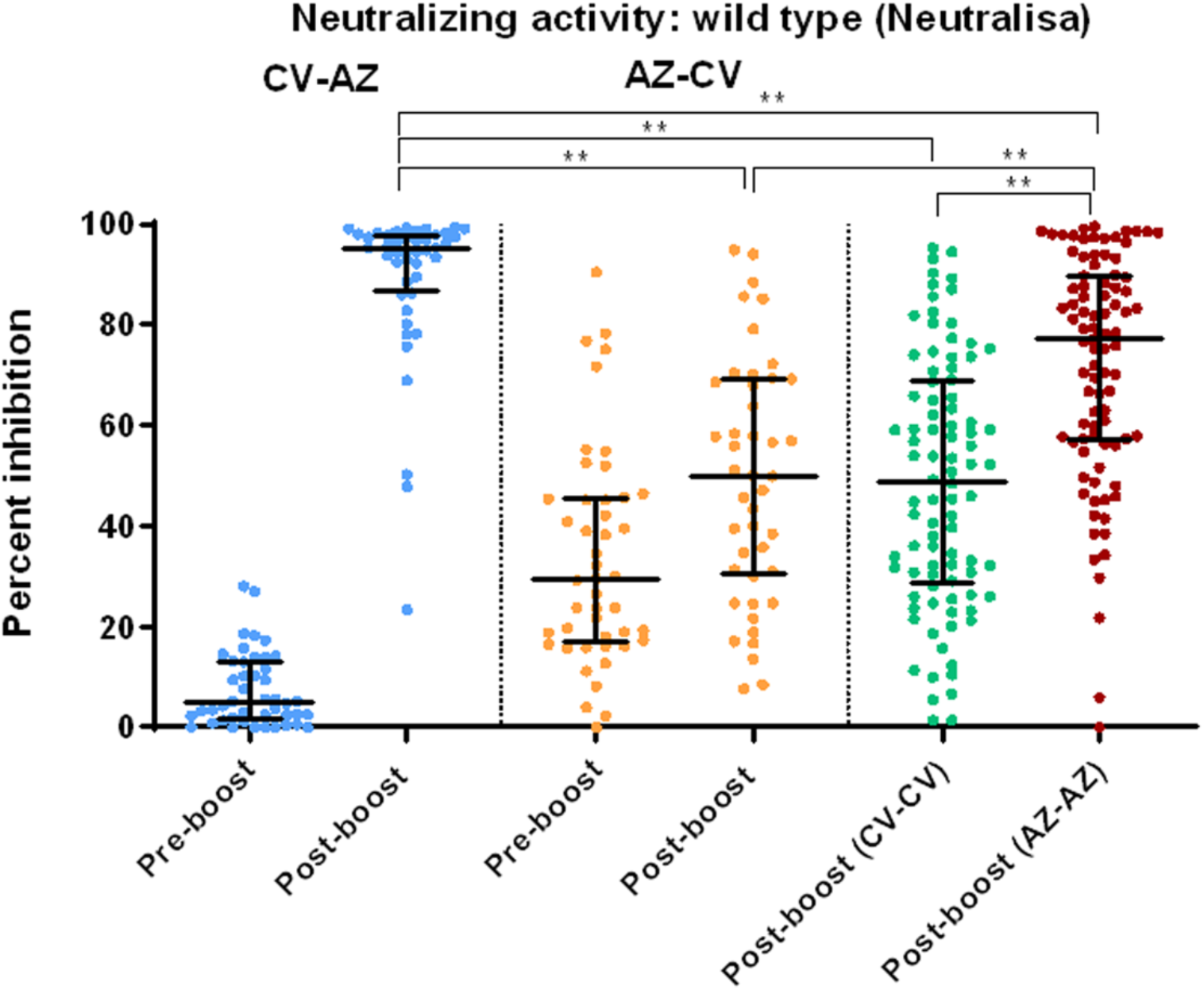
Serum neutralizing activities against wild type SARS-CoV-2 using the surrogate viral neutralization test (sVNT) by Euroimmun (NeutraLISA) in heterologous prime-boost CV-AZ and AZ-CV groups at 4 and 10 weeks after first dose vaccination, respectively (pre-boost), and at 4 weeks after completion of two doses (post-boost). The data points are the reciprocals of the individual. Lines indicate median and bars indicate interquartile ranges. As a reference, serum neutralizing activities were compared 4 weeks after completion of two doses (post-boost) among homologous CV and AZ vaccinees. ** indicates *p* < 0.001.

A comparison of neutralizing activities against wild-type and variant strains one month after completion of two doses showed that the CV-AZ and AZ-AZ groups generated higher neutralizing activities against wild-type and all variant strains (Figure 5) than the AZ-CV and CV-CV group (p < 0.001).

**Figure 5.**
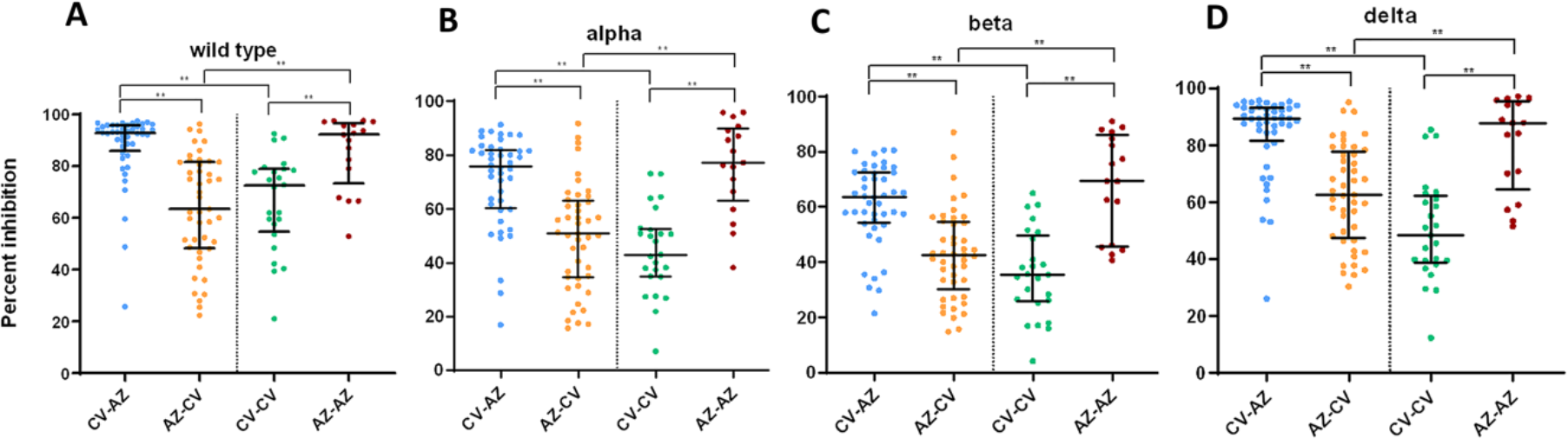
Serum neutralizing activities against wild-type SARS-CoV2 and variants of concern (alpha, beta, and delta) in heterologous and homologous vaccine recipients 1 month after completion of two doses (after boost). The data points are the reciprocals of the individual. Lines indicate median and I-bars indicate interquartile ranges. ** indicates *p* < 0.001.

### Total T cell responses after heterologous vaccinations

Among heterologous CV-AZ vaccine recipients, the IFN-γ responses were observed in at least one Ag tube after second dose vaccination in the majority of subjects (36/44; 82% for Ag 1 and 42/44: 95% for Ag 2). The subtracted IFN-γ response increased significantly increased at post-boost compared to the pre-boost levels (Figure 6A). For heterologous AZ-CV vaccine recipients, we also observed an increase in the subtracted IFN-γ response after booster (Figure 6B), but only 75 % (34/45) and 64% (29/45) of subjects showed increased IFN-γ responses to Ag 1 and Ag 2, respectively. The CV-AZ group elicited a stronger subtracted IFN-γ response after second dose vaccination.

**Figure 6.**
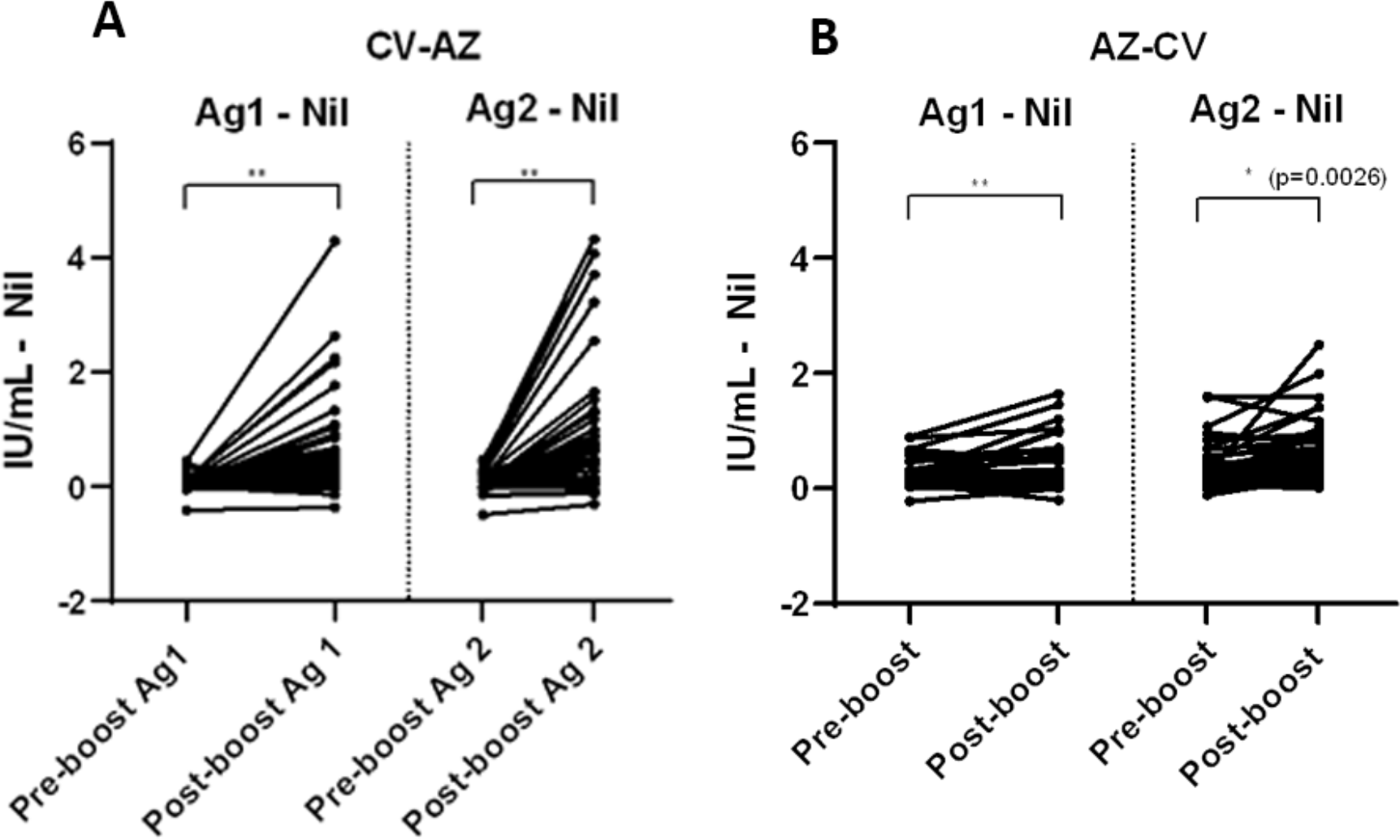
Comparison of subtracted IFN-γ responses using QFN SARS-CoV-2 antigen specific to CD4+ T cells (Ag 1) and CD4+ plus CD8+ T cells (Ag 2) in A) heterologous CV-AZ vaccines and B) heterologous AZ-CV vaccinees at pre-boost and post-boost. A two-tailed pair-matched comparison was performed using the Wilcoxon signed-rank test to analyze the significant differences. ** indicated *p* < 0.001.

## Discussion

Robust data on the safety and immunogenicity of heterologous vaccine regimens will guide the use of these schedules in individuals experiencing AEs after vaccination and in countries experiencing vaccine shortages. Additionally, a mixed vaccination regimen might induce an enhanced immune response compared to homologous licensed schedules. Our study found that heterologous vaccination with CV-AZ induced higher SARS-CoV-2 RBD-specific antibody responses and neutralizing activities against wild type and variants of concern than that of the licensed CV-CV vaccine schedule with proven 65–83% efficacy against symptomatic COVID-19.^16-17^ Furthermore, the CV-AZ schedule, which is administered with a 28-day interval, induced similar RBD-specific binding and neutralizing antibody responses to the licensed AZ-AZ vaccine, which is administered 10–12 weeks apart. Although the cutoff antibody titer that correlates with clinical protection has not yet been clearly defined, a study of nonhuman primate animals has shown that high levels of binding and neutralizing antibodies correlate with the reduction of viral replication in the upper and lower airways after SARS-CoV-2 challenge.^18^ Furthermore, a previous study in humans found that anti-RBD IgG levels of 506 BAU/ml correlated with protection against symptomatic SARS-CoV-2 infection against B.1.1.7 (alpha variant).^19^ The CV-AZ regimen could achieve more than 506 BAU/mL similar to that induced by the AZ-AZ regimen but within a shorter time period, suggesting that the heterologous CV-AZ regimen could be used as an alternative to the proven efficacy of the CV-CV and AZ-AZ regimen in the general population.

Enhanced anti-SARS-CoV-2 spike IgG responses after heterologous vaccination have been reported in ChAdOx1-S/ BNT162b2 (Pfizer-BioNTech) at 28-day^5^ and 73-day intervals^20^ compared to the homologous ChAdOx1-S vaccine. Another small study in healthcare workers also showed higher neutralizing activities in sera of ChAdOx1-S/BNT162b2 recipients than in the sera of homologous ChAdOx1-S and BNT162b2 recipients.^21^ In terms of immune responses against variants, a previous study found that in individuals with ChAdOx1-S-primed individuals, a ChAdOx1-S boost did not induce potent neutralizing antibodies against the delta variant compared to a heterologous boost with an mRNA vaccine.^6^

Nonetheless, not all heterologous regimens result in better immunogenicity compared to the homologous regimen. For example, heterologous vaccination with BNT162b2 followed by ChAdOx1-S did not show enhanced SARS-CoV-2 specific antibody responses compared to homologous BNT162b2.^5^ In the present study, ChAdOx1-S followed by CoronaVac induced lower SARS-CoV-2 RBD-specific antibody responses and neutralizing activities against wild-type and variants of concerns compared to the homologous ChAdOx1-S regimen. When the 2 vaccines are profoundly different, for example, ChAdOx1-S formulation is based on a chimpanzee adenovirus, and CoronaVac makes their vaccine with inactivated whole virion, they induced protective immunity in different mechanisms, possibly resulting in enhanced response compared to the homologous regimen. According to our observations, the whole virion-inactivated vaccine appears to prime the immune system with hundreds of different antigens similar to those elicited by natural infection. This priming could be beneficial in enhancing the subsequent responses by different types of vaccination. However, the inactivated vaccine itself appears not to be potent when used as a booster dose. Further research is needed to elaborate on the immune mechanism induced by whole-virion vaccines.

In this relatively small cohort, the heterologous regimen led to more frequent reports of recorded AE symptoms than the homologous regimen. The reported AE in the homologous groups are in line with what has previously been published.^16,22^ A previous report showed that heterologous schedules incorporating ChAdOx1-S and BNT162b2 vaccines are more reactogenic than the homologous schedule.^15^ This could be explained by the different types of vaccines that induced different types of local inflammation and systemic reactions.

The nucleocapsid protein (N) is abundant in the SARS-CoV-2 virion. The N protein is highly immunogenic and represents a powerful diagnostic and prophylactic target.^23^ In individuals that have recovered from COVID-19, a specific antibody against the N protein can be detected for several months.^24^ Our study showed that administration of the two-dose CoronaVac vaccine containing N protein induced an anti-N IgG response similar to that induced by natural infection. One dose of CoronaVac did not show significant anti-N IgG responses and ChAdOx1-S which contains no N protein did not show any significant anti-N IgG responses. Immunization studies of mice, rats, and nonhuman primates with CoronaVac also showed a specific antinucleocapsid antibody response, although not as much as antibodies against the S protein and the S1-RBD.^25^ The exact role of the nucleocapsid antibody in clinical protection has yet to be identified.

The presence of serum anti-SARS-CoV-2 IgA was documented after natural infection and vaccination with some COVID-19 vaccine platforms such as an mRNA vaccine.^26-28^ Our study showed that in addition to IgG, heterologous prime-boost CV-AZ vaccines also elicited spike-specific IgA, which may be important in preventing transmission and infection. Other combinations including CV-CV, AZ-AZ, and AZ-CV did not induce significant serum IgA responses after two-dose vaccination; however, our earlier observation showed that after the third dose vaccination with AZ in two-dose CV completion, serum IgA could be detected.^29^ Recent studies have found that serum IgA plays a crucial role in virus neutralization.^30^ Serum IgA may reach mucosal surfaces by transduction or through plasmablasts secreting recirculating IgA with a mucosal homing profile.

Interferon gamma responses have been reported after two-dose CoronaVac vaccination, BNT162b1 and recombinant adenovirus type-5 vectored COVID-19.^31-33^ However, there were no standardized techniques to compare the magnitude of the responses between these studies. A previous study also demonstrated a trend of superior T cell responses in heterologous individuals.^5^ The results of this study also showed increased interferon gamma responses after *ex vivo* cell stimulation with SARS-CoV-2 antigens after heterologous CV-AZ and AZ-CV vaccination, suggesting a Th1 immune response that could lead to virus clearance. Notably, the IFN-γ responses were higher in the CV-AZ group than in the AZ-CV group, corresponding to the magnitude of SARS-CoV-2 specific antibody responses in this present study. This T cell together with the antibody responses induced by the heterologous vaccine schedules suggest that it has the potential to protect against COVID-19 through both cellular and humoral immunity. Regarding the timing of the peak T cell response in vaccinated individuals, a previous study showed that the QFN SARS-CoV-2 response peaked at 11–14 days and decreased slightly at 28–32 days.^34^ Therefore, this study could have missed the peaked the IFN-γ responses. Furthermore, previous studies also showed that sustained T cell responses can be detected several months after COVID-19 infection and vaccination^35^ and may last up to several years, as demonstrated in the SARS-CoV-1 study^.36^

Although robust humoral and cellular immune responses are observed following the heterologous schedule, the durability of immune responses and clinical efficacy needs to be further investigated. A previous study showed that immune responses induced by the two-dose CoronaVac vaccine were short-lived^37^, and a third dose vaccination is warranted to increase protection against emerging SARS-CoV-2 variants. Thailand has implemented CV-AZ vaccination in healthy Thai individuals since July 2021, with more than 1.5 million people vaccinated with this heterologous regimen as of 2 September 2021.^38^ Long-term follow-up to monitor immune responses and SARS-CoV-2 infection rates in heterologous CV-AZ cohorts is ongoing to determine the need for the booster dose. Additionally, the adverse event rates observed in this study are subject to variability due to different data collection methods. Furthermore, the interferon gamma release assay was not performed in all vaccinated groups, and other T cell function tests for other cytokines were not performed. The sample size in the present study is limited. Additional studies on a larger group of heterologous vaccinated individuals will be necessary to determine whether the data reflect the general population or whether there are differences due to genetic or environmental factors.

In low- and middle-income countries experiencing a vaccine shortage and emerging variants, heterologous COVID-19 vaccine schedules have the potential to accelerate vaccine rollout. Two-dose vaccination administered in a short time could rapidly increase protective immunity within the population in the middle of the COVID-19 pandemic with emerging variants. Our study demonstrated that heterologous CoronaVac followed by ChAdOx1-S can be considered an alternative regimen to homologous ChAdOx1-S, as it can induce SARS-CoV-2 RBD-specific antibodies and neutralizing activities against wild-type and variants of concerns similar to the licensed two-dose ChAdOx1-S. More studies on the clinical efficacy and durability of immune responses induced by heterologous vaccine regimens are warranted.

## Data Availability

All data produced in the present work are contained in the manuscript.

## Acknowledgements

We would like to thank Prof. Stephen Kerr from the Research Affairs, Faculty of Medicine, Chulalongkorn University for the statistical analysis.

