## Supplementary Information for "Safety and immunogenicity of heterologous and homologous inactivated and adenoviral-vectored COVID-19 vaccines in healthy adults"

**Supplementary Table 1.** GMT (with 95% confidence intervals) of RBD-specific total Ig and anti-RBD IgG, median (with interquartile ranges) of anti-N IgG S/C index, anti-S IgA OD/CO ratios, percentage inhibition against wild-type and variants of SARS-CoV-2 and interferon gamma responses in serum from vaccinated individuals who received homologous CoronaVac (CV) or ChAdOx1-S AstraZeneca (AZ) vaccines and heterologous CV-AZ and AZ-CV vaccines.

|  | **CoronaVac/AZD1222**  **(CV-AZ)** | **AZD1222/CoronaVac**  **(AZ-CV)** | **CoronaVac/CoronaVac**  **(CV-CV)** | **AZD1222/AZD1222**  **(AZ-AZ)** |
| --- | --- | --- | --- | --- |
| **RBD-specific total Ig (U/mL)** | | | | |
| Pre-prime  GMT (95% CI) | 0.4 (0.4-0.4) | 0.4 (0.4-0.4) | N/A | N/A |
| n | 46 | 48 | N/A | N/A |
| Pre-boost  GMT (95% CI) | 2.2 (1.5-3.1) | 65.3 (48.2-88.4) | N/A | N/A |
| n | 46 | 46 | N/A | N/A |
| Post-boost  GMT (95% CI) | 573.4 (417.2-788.0) | 375.8 (304.9-463.3) | 99.4 (77.5-127.3) | 933.7 (775.4-1124.0) |
| n | 44 | 46 | 90 | 89 |
| **Anti-RBD IgG (BAU/mL)** | | | | |
| Pre-prime  GMT (95% CI) | 1.1 (0.9-1.2) | 1.0 (0.9-1.1) | N/A | N/A |
| n | 46 | 48 | N/A | N/A |
| Pre-boost  GMT (95% CI) | 9.4 (7.0-12.7) | 51.6 (39.2-67.8) | N/A | N/A |
| n | 46 | 46 | N/A | N/A |
| Post-boost  GMT (95% CI) | 664.8 (520.0-850.0) | 88.1 (71.9-107.9) | 116.2 (99.5-135.7) | 1297.0 (1075.0-1566.0) |
| n | 44 | 46 | 90 | 89 |
| **Anti-N IgG (S/C index)** | | | | |
| Pre-prime  Median (IQR) | 0.02 (0.02-0.03) | 0.02 (0.02-0.04) | N/A | N/A |
| n | 46 | 48 | N/A | N/A |
| Pre-boost  Median (IQR) | 0.04 (0.03-0.07) | 0.03 (0.02-0.04) | N/A | N/A |
| n | 46 | 46 | N/A | N/A |
| Post-boost  Median (IQR) | 0.05 (0.03-0.07) | 0.08 (0.05-0.15) | 0.8 (0.3-2.1) | 0.03 (0.02-0.07) |
| n | 44 | 46 | 90 | 89 |
| **Anti-S IgA (OD/CO)** | | | | |
| Pre-prime  Median (IQR) | 0.3 (0.2-0.3) | 0.2 (0.1-0.4) | N/A | N/A |
| n | 15 | 15 | N/A | N/A |
| Pre-boost  Median (IQR) | 0.6 (0.4-1.0) | 0.6 (0.4-1.5) | N/A | N/A |
| n | 45 | 45 | N/A | N/A |
| Post-boost  Median (IQR) | 4.7 (2.9-7.9) | 0.8 (0.5-1.7) | 1.0 (0.6-2.4) | 1.2 (0.6-2.1) |
| n | 42 | 44 | 30 | 30 |
| **Neutralizing activities (percent inhibition)** | | | | |
| **sVNT**—**Wild Type**  (Euroimmun, Lubeck, Germany)  Pre-boost  median (IQR) | 4.9 (1.7-13.1) | 29.4 (17.1-45.4) | N/A | N/A |
| **n** | 45 | 46 | N/A | N/A |
| **sVNT**—**Wild Type**  (Euroimmun, Lubeck, Germany)  Post-boost  median (IQR) | 95.0 (86.8-97.8) | 50.0 (30.3-69.0) | 48.8 (28.9-68.8) | 77.2 (57.0-89.7) |
| **n** | 44 | 44 | 90 | 89 |
| **sVNT**—**Wild Type**  **(**GenScript, Jiangsu, China**)**  Post-boost  median (IQR) | 92.8 (85.9-95.7) | 63.5 (48.4-81.5) | 72.4 (54.7-78.8) | 92.1 (73.5-96.6) |
| **n** | 44 | 45 | 24 | 17 |
| **sVNT—B.1.1.7 (alpha)**  **(**GenScript, Jiangsu, China**)**  Post-boost  median (IQR) | 75.7 (60.3-82.0) | 50.8 (34.5-63.1) | 43.1 (35.0-52.6) | 77.2 (63.1-90.0) |
| n | 44 | 45 | 25 | 17 |
| **sVNT—B.1.351 (beta)**  **(**GenScript, Jiangsu, China**)**  Post-boost  median (IQR) | 63.6 (54.2-72.5) | 42.7 (30.1-54.4) | 35.4 (25.8-49.7) | 69.3 (45.7-85.9) |
| n | 44 | 45 | 25 | 17 |
| **sVNT—B.1.617.2 (delta)**  **(**GenScript, Jiangsu, China**)**  Post-boost  median (IQR) | 89.3 (81.7-93.1) | 62.6 (47.4-77.6) | 48.4 (38.9-62.2) | 87.7 (64.6-95.3) |
| n | 44 | 45 | 25 | 17 |
| **Interferon gamma release assay (IU/mL minus Nil)** | | | | |
| IFN-γ Ag 1  Pre-boost | 0.03 (0.00-0.06) | 0.11 (0.05-0.24) | N/A | N/A |
| n | 46 | 46 | N/A | N/A |
| IFN- γ Ag 1  Post-boost | 0.30 (0.10-0.80) | 0.21 (0.06-0.56) | N/A | N/A |
| n | 44 | 45 | N/A | N/A |
| IFN- γ Ag 2  Pre-boost | 0.01 (0.00-0.08) | 0.23 (0.08-0.45) | N/A | N/A |
| n | 46 | 46 | N/A | N/A |
| IFN- γ Ag 2  Post-boost | 0.65 (0.14-1.26) | 0.28 (0.11-0.83) | N/A | N/A |
| n | 44 | 45 | N/A | N/A |

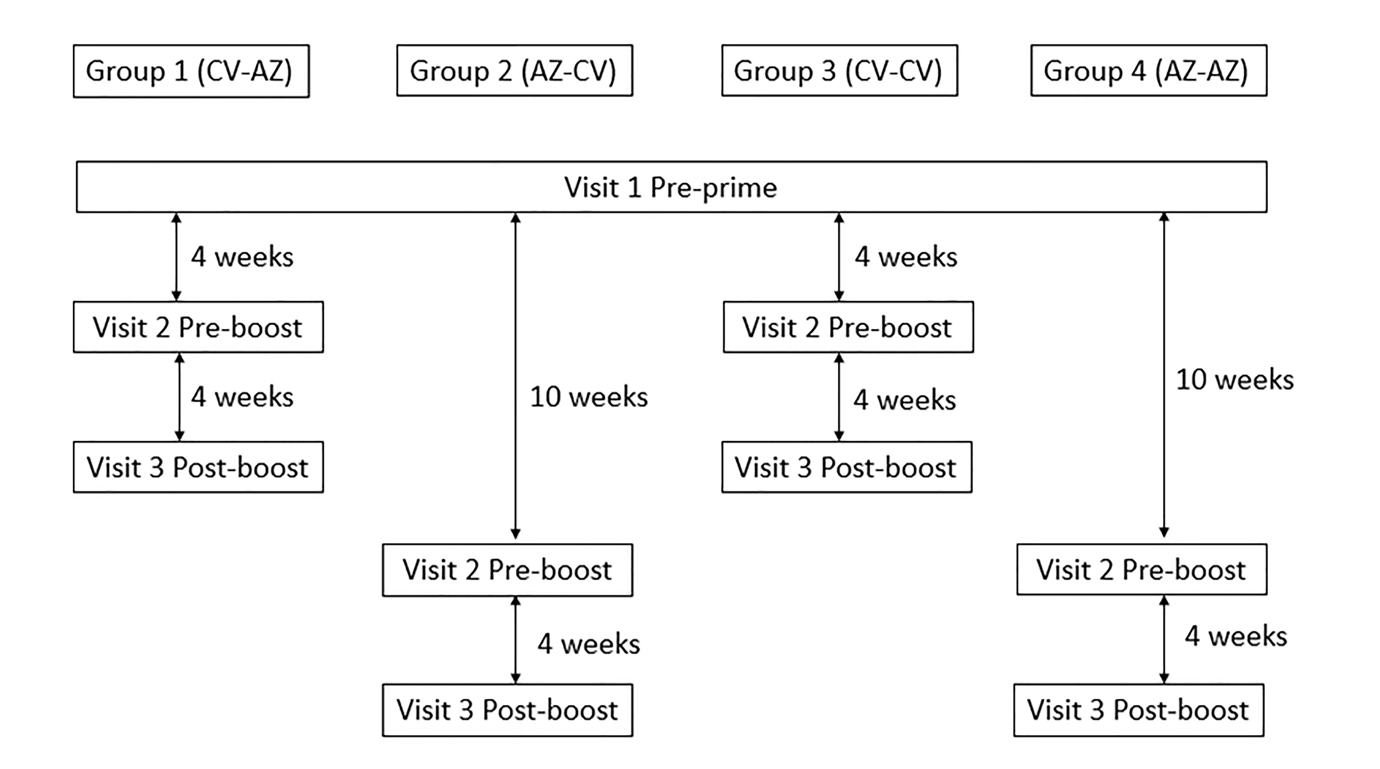

**Figure S1.** Participant flow chart in homologous and heterologous cohorts.

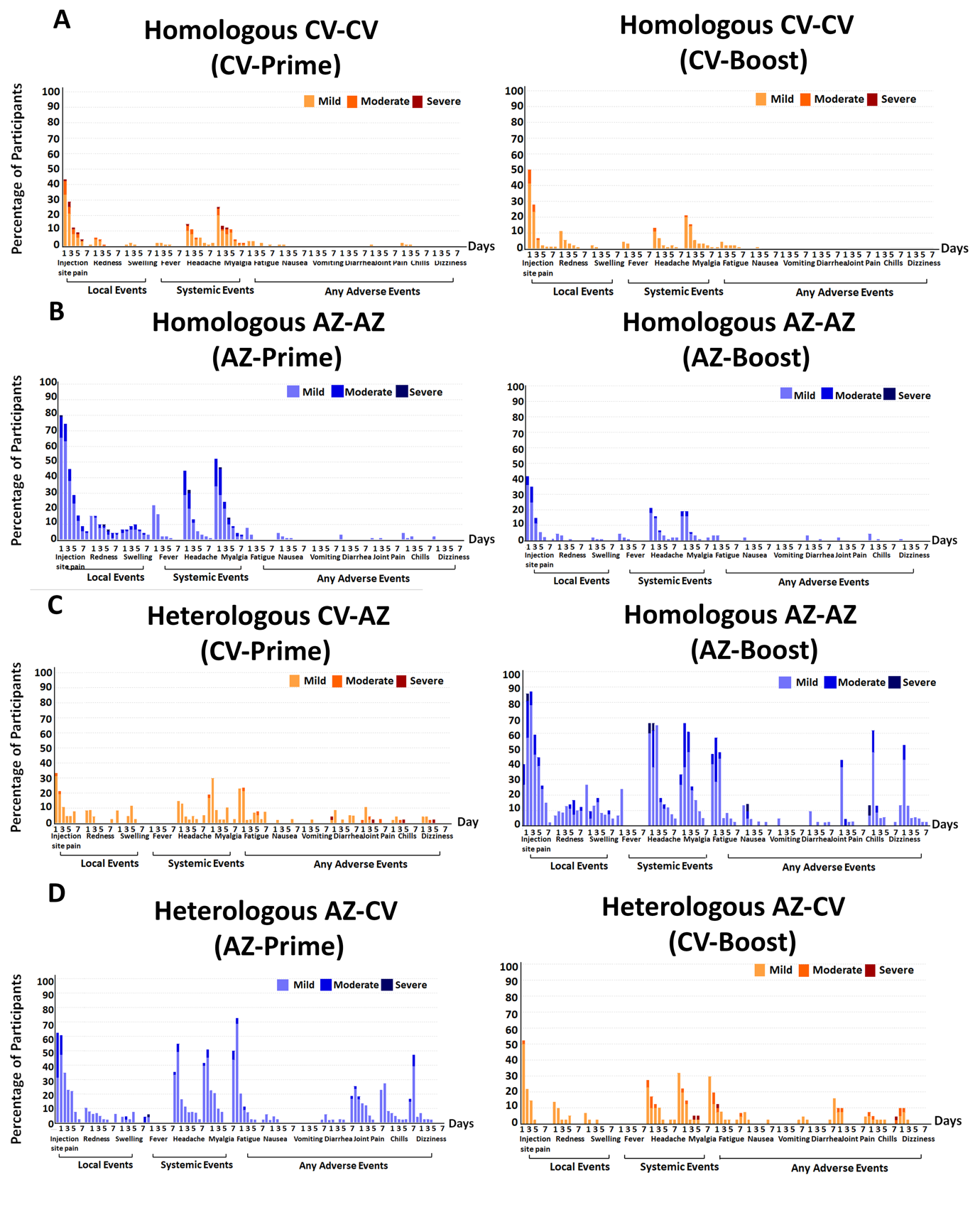

**Figure S2.** Adverse events (AEs) within 7 days after the prime and booster dose after A) homologous CV-CV vaccination B) homologous AZ-AZ vaccination C) heterologous CV-AZ vaccination and D) heterologous AZ-CV vaccination. The percentage of participants who reported local and systemic AEs is shown on the y-axis. Fever is defined as mild: 38·0°C to <38.5°C; moderate: 38.5°C to <39°C; severe: ≥39.0°C. For systemic symptoms, the classification was follows: mild–easily tolerated with no limitation in normal activity; moderate–some limitation in daily activity; severe–unable to perform normal daily activity.^15^
